# Personalized Inhaled Bacteriophage Therapy Decreases Multidrug-Resistant *Pseudomonas aeruginosa*

**DOI:** 10.1101/2023.01.23.22283996

**Authors:** BK Chan, GL Stanley, KE Kortright, M Modak, IM Ott, Y Sun, S Würstle, C Grun, B Kazmierczak, G Rajagopalan, Z Harris, CJ Britto, J Stewart, JS Talwalkar, C Appell, N Chaudary, SK Jagpal, R Jain, A Kanu, BS Quon, JM Reynolds, QA Mai, V Shabanova, PE Turner, JL Koff

## Abstract

Bacteriophage therapy, which uses lytic viruses as antimicrobials, has received renewed interest to address the emerging antimicrobial resistance (AMR) crisis. Cystic fibrosis (CF), a disease complicated by recurrent *P. aeruginosa* pulmonary infections that cause lung function decline, is an example where AMR is already a clinical problem. While bacteria evolve bacteriophage resistance, we developed a strategy to select bacteriophages that target bacterial cell surface receptors that contribute to antibiotic resistance or virulence. Thus, in addition to killing bacteria, these phages steer surviving, evolved bacteria to antibiotic re-sensitivity or attenuated virulence. Here, we present outcomes from nine CF adults treated with nebulized bacteriophage therapy for AMR *P. aeruginosa* using this personalized approach. Results showed that phage therapy: 1) reduced sputum *P. aeruginosa*, 2) showed evidence for predicted trade-offs in most subjects, and 3) improved lung function, which may reflect the combined effects of decreased bacterial sputum density and phage-driven evolved trade-offs.

## Introduction

The rise of antimicrobial resistance (AMR) poses a serious global threat to human health [1]. By 2050 AMR-induced mortality is projected to overtake annual deaths caused by currently more-common diseases (e.g., cancer and diabetes) due to increasing multidrug-resistant (MDR) and pan-drug-resistant (PDR) bacterial infections [1]. The ESKAPE pathogens (*Enterococcus faecium, Staphylococcus aureus, Klebsiella pneumoniae, Acinetobacter baumannii, Pseudomonas aeruginosa*, and *Enterobacter spp*) are particularly concerning because they can present simultaneous resistance against multiple classes of antibiotics [2]. Complications due to MDR/PDR pathogens are already increasingly common in cystic fibrosis (CF), which is one of the most common life-limiting monogenic diseases. The most prevalent bacterial infections in CF adults are due to the ESKAPE-pathogen *P. aeruginosa* (PsA), which significantly predicts disease morbidity and mortality [3].

Bacteriophage (phage) therapy has received renewed interest as a potential anti-infective to treat MDR/PDR infections [4]. Lytic phages are bacteria-specific viruses that replicate within susceptible bacterial cells, which they lyse (kill) to release progeny phages. Thus, phages can act as self-amplifying, personalized ‘drugs’ that propagate in host bacterial cells within a treated patient [4, 5]. However, bacteria employ a range of defense mechanisms, which can improve through evolution to better resist phage(s) [6]. This is a recognized vulnerability of phage therapy that should limit its clinical utility. However, studies suggest that certain phages can kill target bacteria while selecting for evolved resistance that coincides with a clinically-useful genetic trade-off; here, the bacteria evolve phage-resistance that improves fitness but suffer reduced performance in a pathogenicity trait (e.g., decreased virulence or antibiotic re-sensitization) [4]. Therefore, implementing a phage therapy strategy that purposefully leverages a trade-off could result in improved clinical outcomes by 1) using phages to decrease MDR/PDR bacterial burden, and 2) selecting for surviving bacterial mutants that evolve phage resistance and become less virulent or re-sensitized to antibiotics [7].

We developed an approach to select environmentally sourced lytic phages that are naturally capable of binding to bacterial cell surface receptors that contribute to functional mechanisms of antibiotic resistance (e.g., efflux pumps), or virulence [e.g., lipopolysaccharide (LPS) and type-IV pili (TIVP)] [4]. In addition to killing target bacteria, these phages were chosen for their ability to predictably steer surviving bacterial mutants, by exerting selection for altered (or deleted) binding receptors, which enrich for antibiotic re-sensitivity or attenuated virulence should phage-resistance evolve. For example, phage OMKO1 kills PsA and selects for evolved phage resistance that coincides with reduced multi-drug efflux (Mex) pump function to achieve reduced antibiotic resistance [8], a trade-off exploited in a case of phage therapy [9]. Using this approach, the current study also deployed two additional lytic phages that were found to bind to LPS or to TIVP, which drive evolved phage-resistance trade-offs that compromise bacterial virulence.

Here, we present outcomes from nine adult patients with CF (PwCF) who were consecutively treated with nebulized phage therapy for sputum MDR/PDR PsA using the above personalized approach. Phage therapy was safe, and results showed that phage therapy reduced PsA sputum bacteria, without significantly altering the lung microbiome in favor of other CF pathogens. Our data provided evidence that in most subjects, post-phage therapy sputum PsA showed evidence for predicted trade-offs. Following phage therapy, PwCF showed improvement in lung function, which may reflect the combined effects of decreased bacterial sputum density and phage-driven evolved trade-offs.

## Results

### Treatment Protocol

PwCF sent unsolicited requests for assistance to treat MDR/PDR PsA infections that did not respond to standard CF therapies (Table 1). This consecutive cohort included 8 women and 1 man with CF, median age 32 years (range: 22-46 years). Sputum PsA was PDR (3 patients) or MDR (6 patients), and all patients had a clinical course complicated by frequent pulmonary exacerbations despite oral, inhaled, and/or intravenous (IV) antibiotics. At the time of phage therapy, all patients either recently completed antibiotics or were concurrently on IV antibiotics. Some patients had additional CF pathogens cultured in sputum (e.g., *Staphylococcus aureus, Achromobacter spp*., and non-tuberculous mycobacteria; Table 1). At the time of phage therapy, elexacaftor/tezacaftor/ivacaftor (Trikafta/Kaftrio^®^) was not available. Four individuals were taking tezacaftor/ivacaftor (Symdeko/Symkevi^®^; Table 1).

**Table 1.** Demographics and Clinical Characteristics of Patient Pre-Phage. **Cystic Fibrosis Subject Demographics and Clinical Characteristics**. Demographics and clinical characteristics of nine (9) cystic fibrosis (CF) adults prior to bacteriophage (phage) therapy. CF transmembrane conductance regulator (CFTR) genetic mutation is characterized as delta F508 (delF508) homozygous, heterozygous, or other. Complications associated with CF, range of lung function, number of pulmonary exacerbations (episodes of clinical deterioration associated with respiratory infection) and CF therapies are shown. Number of subjects with multi-drug resistant (MDR) or pan-drug resistant (PDR) *Pseudomonas aeruginosa* are shown. Additional CF sputum pathogens pre-phage therapy are reported. Post-phage therapy there was no change in these pathogens based on clinical laboratory sputum cultures.

| **Table 1. Demographics and Clinical Characteristics of Patient Pre-Phage** | **Patients (N=9)** |
| --- | --- |
| **Age** |  |
| 18-30 | 6 (67%) |
| > 30 | 3 (33%) |
| **Sex** |  |
| Female/Male | 8 (89%)/1 (11%) |
| **CFTR Genotype** |  |
| F508del/F508del | 3 (33%) |
| F508del/Other* | 4 (44%) |
| Other*/Unknown | 2 (22%) |
| **CF Complications** |  |
| Pancreatic Insufficiency | 9 (100%) |
| Malnutrition (BMI: < 22 female or < 23 male) | 8 (89%) |
| CF-Related Diabetes | 6 (67%) |
| Allergic Bronchopulmonary Aspergillosis | 2 (22%) |
| **FEV<sub>1</sub>%** |  |
| 30-69% | 5 (56%) |
| < 30% | 4 (44%) |
| **Pulmonary Exacerbations** |  |
| > 4/year | 9 (100%) |
| **CF Therapies** |  |
| CFTR Modulator |  |
| Tezacaftor/ivacaftor | 4 (44%) |
| Elexacaftor/tezacaftor/ivacaftor | 0 |
| Mucolytics (rDNase, hypertonic saline) | 9 (100%) |
| Systemic steroids | 3 (33%) |
| Inhaled antibiotics | 9 (100%) |
| IV antibiotics during phage therapy | 7 (78%) |
| ***Pseudomonas aeruginosa* Antibiotic Resistance** |  |
| MDR | 6 (67%) |
| PDR | 3 (33%) |
| **Other Pathogens<sup>†</sup>** |  |
| *Staphylococcus aureus* | 3 (33%) |
| *Achromobacter* | 2 (22%) |
| *Stenotrophomonas, Burkholderia* | 0 |
| *Mycobacterium avium complex* | 1 (11%) |
| *Mycobacterium abscessus* | 1 (11%) |

\*V520F, 621+1G>T, CFTRdele2,3, G85E, Y913X, 3659delC, 3959delC, 3850-3T>G, R75Q
<sup>†</sup>No changes in other pathogens after phage therapy

Personalized phage therapy was chosen based on sputum MDR/PDR PsA phage susceptibility (Table 2). After suitable phage(s) were identified, a phage therapy protocol was reviewed with each subject’s CF physician, and this protocol was approved *via* individual U. S. Food and Drug Administration (FDA) emergency investigational new drug (eIND) and Institutional Review Board. Nebulization was chosen because several CF therapies are nebulized, such therapies are generally well-tolerated, and phage nebulization may reduce systemic toxicity. Subjects were treated at their CF program (n = 4) or traveled to Yale (n = 5). The Yale Center for Phage Biology & Therapy prepared and provided phages for all subjects.

**Table 2.**
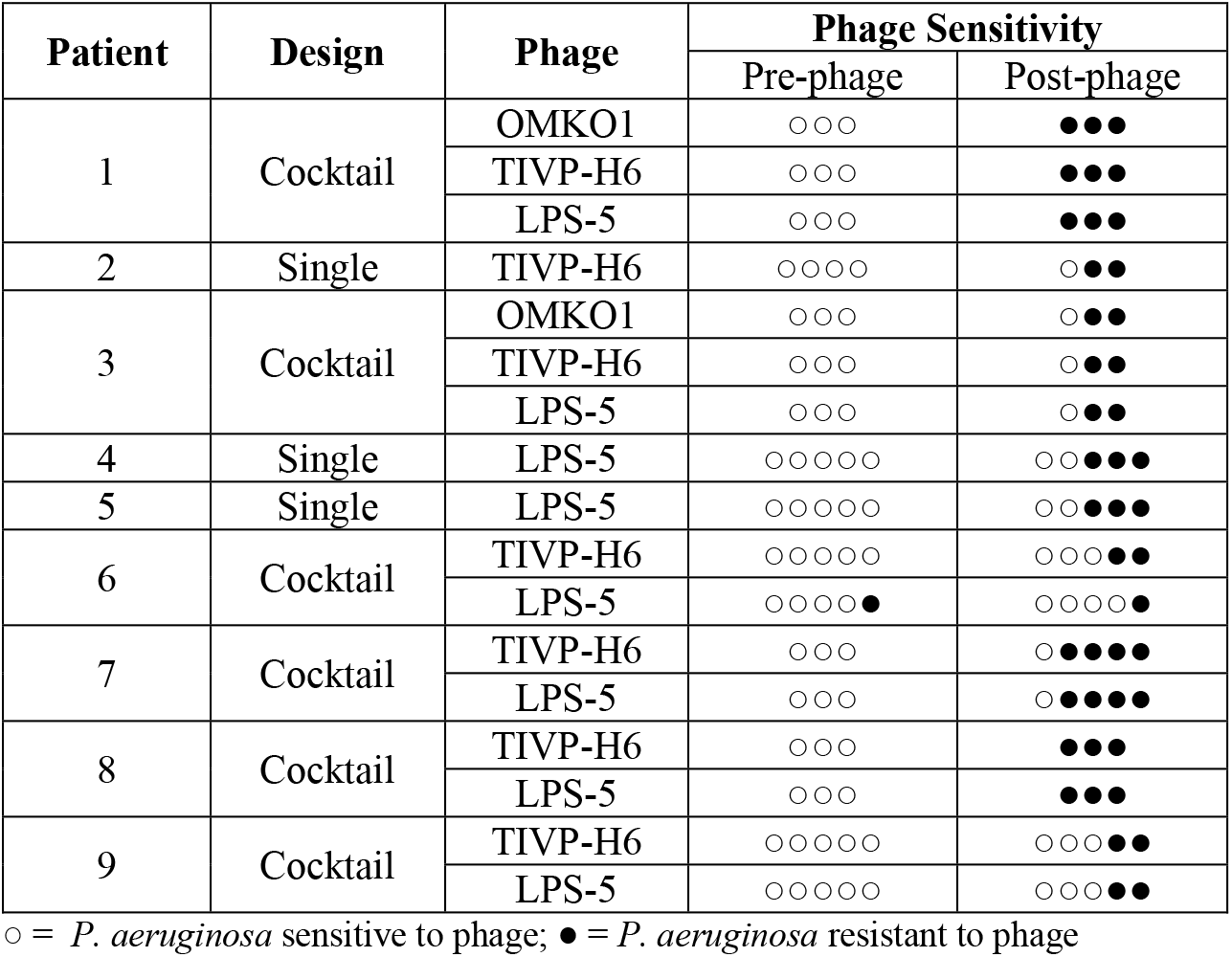
Phage Therapy. **Selection of Phage Therapy and *Pseudomonas* Phage Resistance**. Phages OMKO1, TIVP-H6, and LPS-5 were evaluated, and phages selected for therapy are shown (Column 3) for each subject. MDR/PDR *Pseudomonas aeruginosa* isolate sensitivity for each phage prior to phage therapy (Column 4), and after phage therapy (Column 5), was measured. Empty circles, *Pseudomonas aeruginosa* is sensitive to indicated phage(s); filled circles *Pseudomonas aeruginosa* is resistant to indicated phage(s).

Treatment with inhaled phage therapy occurred twice daily for inpatients (4 subjects) or daily for outpatients (5 subjects; 3 subjects transitioned from inpatient to outpatient) for 7 to 10 days. Inpatients were provided jet nebulizers; outpatients used their own jet nebulizers. Phages were delivered as mixtures (cocktails) of two or three phages (6 subjects) or single phage therapy (3 subjects; Table 2), administered in identical total PFU/ml. No adverse events were attributed to phage nebulization. Several subjects reported low grade temperatures and fatigue that typically occurred from day 2 to 4 of phage therapy. No additional treatment was required for these symptoms. To look for evidence of phage-driven phenotypic trade-offs, sputum was collected and PsA was studied pre- and post-phage treatment. Spirometry was obtained pre- and post-phage treatment.

### Inhaled Phage Therapy Decreases Sputum *Pseudomonas*

MDR/PDR PsA infections pose an increasing therapeutic challenge in CF, which may be amenable to phage therapy. Therefore, spontaneously expectorated sputum collected pre- and post-phage therapy was processed for PsA quantification to test the hypothesis that phage therapy would reduce sputum PsA in each subject. Results showed that for all subjects, sputum PsA pre-phage was greater than post-phage (Fig. 1A). Across all subjects, sputum PsA decreased following phage therapy from a median (1^st^ quartile, 3^rd^ quartile) of 2.6 × 10^8^ (5.7 × 10^8^, 5.0 × 10^7^) or mean of 3.0 × 10^8^ (*±* 1.0 × 10^8^ SEM) CFU/ml before therapy, to median of 2.4 × 10^4^ (1.3 × 10^4^, 5.5 × 10^5^) or mean of 7.0 × 10^6^ (*±* 6.9 × 10^6^ SEM) CFU/ml after therapy, which is a 4 log median, or 2 log mean difference, respectively (p = 0.004, Wilcoxon signed rank t test). Thus, sputum PsA decreased post-phage therapy in each patient and across the cohort, regardless of individualized differences in the administered inhaled phage(s) and treatment strategies (Table 2).

**Figure 1.**
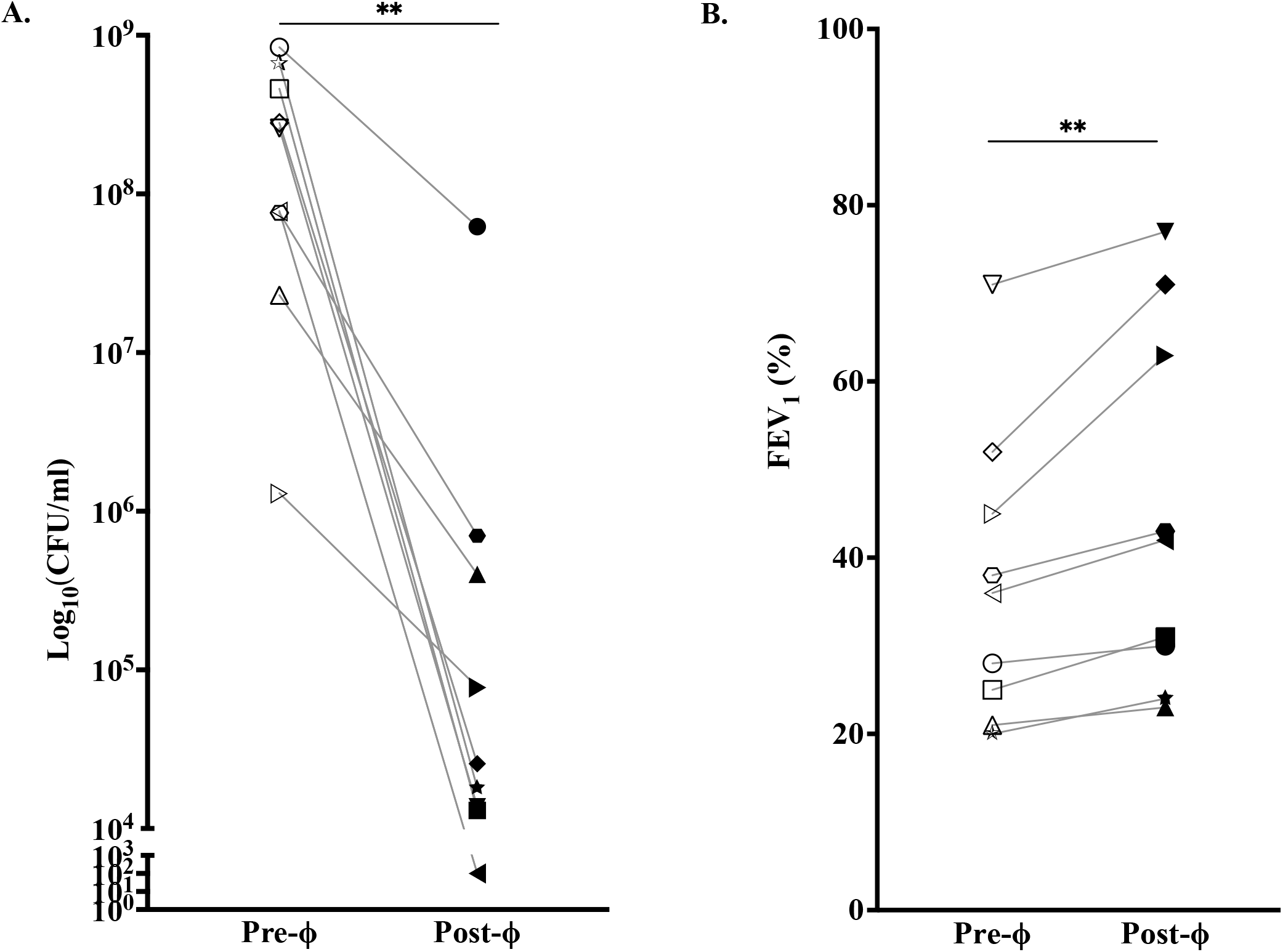
Effect of Phage Therapy on *Pseudomonas* and Lung Function. (**A)**. <u>Effect of Phage Therapy on Sputum</u> *<u>Pseudomonas</u>*. Sputum analysis was performed before and after phage therapy. *Pseudomonas aeruginosa* CFU/ml from each Subject sputum (n = 9) were counted in replicates of 3 and averaged. Pre-phage therapy (Pre-ϕ) CFU/ml median (1^st^ quartile, 3^rd^ quartile) of 2.6 × 10^8^ (5.7 × 10^8^, 5.0 × 10^7^) or mean of 3.0 × 10^8^ (*±* 1.0 × 10^8^ SEM) was measured. Post-phage therapy (Post-ϕ) CFU/ml median of 2.4 × 10^4^ (1.3 × 10^4^, 5.5 × 10^5^) or mean of 7.0 x10^6^ (*±* 6.9 × 10^6^ SEM) CFU/ml was measured, a median 4 log difference or mean 2 log difference, respectively (**p = 0.004, Wilcoxon signed rank t test). (**B)**. <u>Effect of Phage Therapy on Lung Function</u>. Spirometry was performed before and after phage therapy. Precent predicted forced expiratory volume in 1 second (ppFEV_1_) Pre-phage therapy (Pre-ϕ) median (1^st^ quartile, 3^rd^ quartile) of 36 (23, 49) or mean of 37 *±* 5.5 SEM improved to Post-phage therapy (Post-ϕ) median of 42 (27, 67) or mean of 45 *±* 6.9 SEM, a median difference of 6, or mean difference of 8, respectively (**p = 0.004, Wilcoxon signed rank t test).

### Inhaled Phage Therapy Improves Lung Function

In CF, chronic PsA is associated with increased morbidity and mortality due to recurrent pulmonary exacerbations and lung function decline over time. Therapies that decrease PsA have been associated with clinical benefits. Based on observed declines in sputum PsA post-phage treatment (Fig. 1A), we compared lung function [percent predicted FEV_1_ (ppFEV_1_)] pre- to post-phage (day 21 to 35) therapy. Results for each subject showed an increase in ppFEV_1_ post-versus pre-phage therapy (Fig. 1B), and an analysis across the entire cohort showed an overall improvement in ppFEV_1_ from a median (1^st^ quartile, 3^rd^ quartile) of 36 (23, 49), or mean of 37 *±* 5.5 SEM, before therapy to a median of 42 (27, 67), or mean of 45 *±* 6.9 SEM, after therapy, which is median or mean difference in ppFEV1 of approximately 6 and 8, respectively (p = 0.004, Wilcoxon signed rank t test). Analysis of subjects with pre-phage ppFEV_1_ >30 showed increased ppFEV_1_ after phage therapy [mean difference of 11 *±* 3.2 SEM (p = 0.06)], while subjects with pre-phage ppFEV_1_ <30 improved ppFEV_1_ to a lesser extent [mean difference 3.5 *±* 0.96 SEM (p = 0.1)]. Therefore, personalized inhaled phage therapy provided a clinical benefit as measured by improved lung function, regardless of differences in treatment strategies administered (Table 2).

### Phage Therapy Reduces *Pseudomonas* Virulence

In addition to bacterial killing (Fig. 1A), we hypothesized that our personalized strategy to use specific phages to target PsA efflux pumps or virulence factors should select for surviving bacterial mutants with evolved phage resistance that results in decreased virulence. First, PsA resistance to phage(s) administered in inhaled phage therapy was investigated. To do so, purified PsA clones from subject sputum were cultured, and each isolate was tested for susceptibility to phage(s) used in inhaled therapy (Table 2). For each subject, results showed that post-phage sputum PsA contained one or more bacterial clones with resistance to phage(s) used in therapy (Table 2). This result showed that inhaled phage therapy exerted selection for PsA to evolve resistance to phage(s) used in treatment.

Next, we examined whether evolved PsA phage resistance post-phage therapy coincided with a specific virulence factor trade-off. PsA deploys efflux pumps to extrude multiple classes of antibiotics that permeate the bacteria cell to develop antibiotic resistance. Phage OMKO1 selects for evolved PsA phage resistance that coincides with increased sensitivity to various antibiotics [8, 9], which can be explained by phage binding to MEX efflux pumps when infecting PsA [8, 9]. In the two subjects that received OMKO1 phage therapy (Table 2), post-phage therapy sputum PsA evaluated in clinical laboratories showed increased antibiotic sensitivity (Fig. 2A). This result suggested that re-sensitivity to multiple antibiotics occurred with OMKO1.

**Figure 2.**
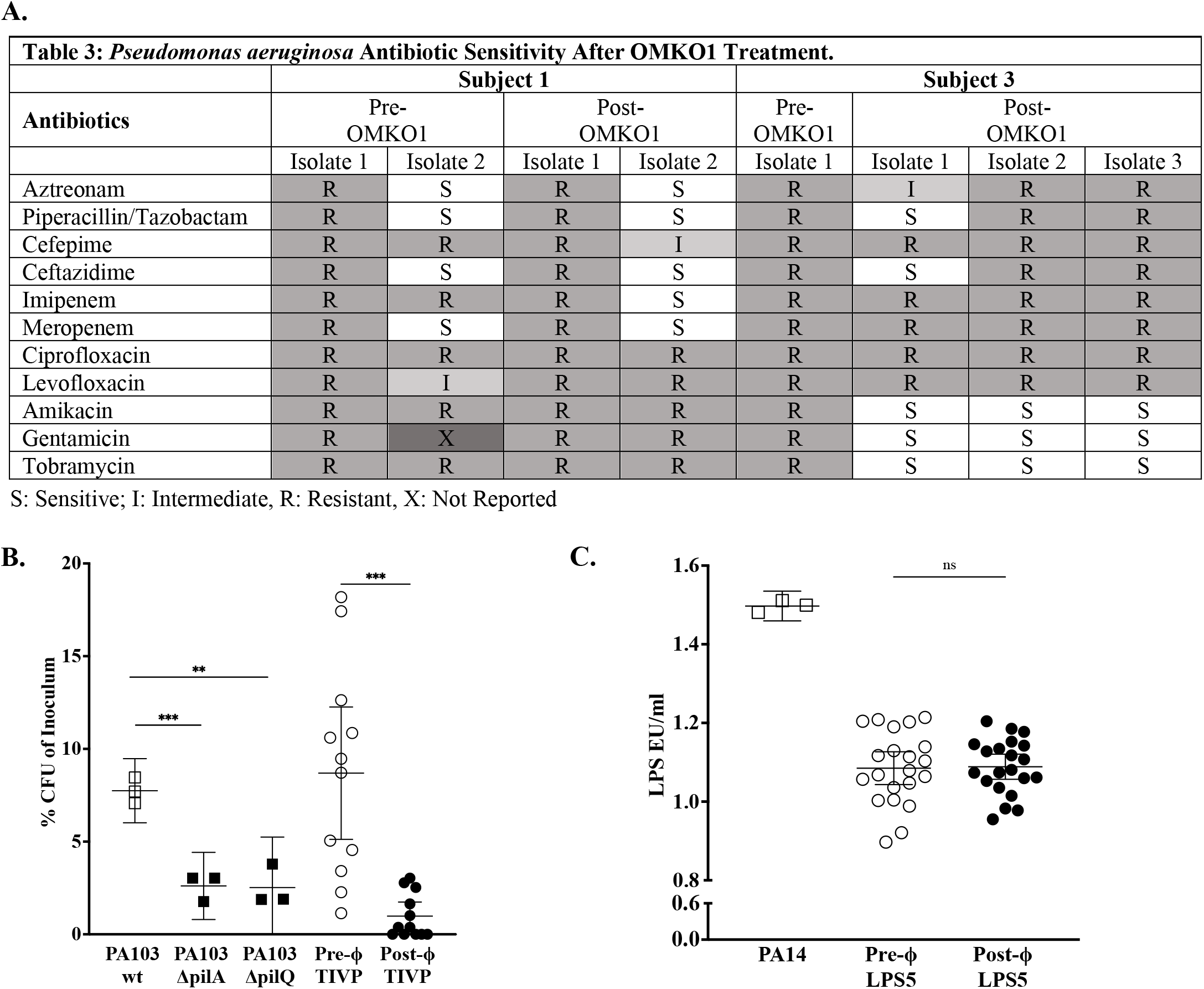
Effect of Phage Therapy on *Pseudomonas* Antibiotic Resistance and Virulence. **(A)**. <u>Effect of Phage Therapy on</u> *<u>Pseudomonas</u>* <u>Antibiotic Resistance</u>. Phage therapy with OMKO1, which targets *Pseudomonas* efflux pumps, results in increased antibiotic sensitivity. In two Subjects (#1 and #3) who received OMKO1 phage therapy, *Pseudomonas aeruginosa* antibiotic sensitivity testing completed from clinical laboratories is shown comparing pre- and post-phage therapy. Subject #1 two isolates reported pre- and post-phage therapy. Subject #3 one isolate pre-phage therapy compared to three isolates post-phage therapy. S = sensitive, I = intermediate, R = resistant, and X = not reported. **(B)**. <u>Effect of Phage Therapy on</u> *<u>Pseudomonas</u>* <u>Virulence</u>. (**B)** *Pseudomonas aeruginosa* attachment to CF airway epithelial (CFBEo-) cells was measured. Bacteria from different *Pseudomonas* laboratory strains wildtype (WT) PA103, PA103 TIVP mutants PA103ΔpilA and PA103ΔpilQ, and four (4) PsA isolates pre-TIVP-H6 phage (Pre-ϕ) and three (3) PsA isolates post-TIVP-H6 phage (Post-ϕ) TIVP from Subject 2 (the only individual who received TIVP-H6 single phage therapy). Each isolate was added to CFBEo-cells at air-liquid interface and attachment was measured in triplicates (CFU percentage of the original inoculum). PA103 WT vs. PA103ΔpilA; N = 1-2, in triplicate (*** p < 0.001; Welch’s t test); PA103 WT vs. PA103ΔpilQ; N=1-2, in triplicate (** p < 0.01; Welch’s t test). Pre-ϕ vs. post-ϕ TIVP phage therapy isolates; N = 3-4, in triplicate (*** p < 0.001; Welch’s t test). (**C)** Endotoxin was measured in *Pseudomonas aeruginosa* supernatants after LPS phage therapy. LPS EU/ml was measured (corrected for CFU/ml) in a *Pseudomonas* laboratory strain (PA14) and pre- and post-LPS phage cell-free PsA supernatants from Subjects 4 and 5 (individuals who received LPS-5 single phage therapy), N = 7, in triplicate (comparison was non-significant; Welch’s t test).

Type-IV pili (TIVP) function in PsA twitching motility and surface attachment to airway epithelium in the CF lung [10]. Phage TIVP-H6 binds to PsA TIVP (data not shown), and the hypothesis that TIVP-H6 phage therapy should select against PsA TIVP production was investigated. Here, pre- and post-phage therapy PsA isolates from a subject that received TIVP-H6 (Table 2) were isolated, and TIVP function was studied in a standard attachment assay on human CF airway epithelial (CFBE41o-) cell line cultured at air-liquid interface *in vitro* (Fig. 2B). Equal amounts of PsA were isolated and evaluated for adherence to CFBE41o-cells. Results also compared adherence of laboratory wildtype PsA strain (PA103) to that of engineered mutants deficient in TIVP production (Δ*pilA* and Δ*pilQ*) and confirmed that decreased adherence of each mutant was statistically significant (Fig. 2B). In addition, post-phage therapy TIVP-H6 PsA isolates showed decreased adherence to CF airway epithelium, compared with pre-phage PsA (Fig. 2B). Thus, these results showed that phage TIVP-H6, which kills PsA (Fig. 1), also selected against TIVP production in phage-resistant bacteria (Table 2 & Fig. 2B).

PsA LPS reduces antibiotic permeability, contributes to biofilm production, and activates human host immune responses via Toll-like receptor 4. In contrast, during chronic infection(s), decreased PsA LPS results in reduced immunostimulatory potential, which enables bacteria to evade the host immune system [11]. Phage LPS-5 binds to PsA LPS (data not shown), and the hypothesis that LPS-5 phage therapy should select against PsA LPS production was studied. Here, equal amounts of PsA were isolated and LPS concentration was measured in pre- and post-phage therapy PsA isolates from two subjects that received phage LPS-5 (Table 2). Results compared endotoxin units (EU/ml) measured for supernatants from wildtype (PA14) and PsA pre- and post-LPS-5 phage therapy (Fig. 2C). PsA pre- and post-LPS-5 phage therapy did not show a difference in LPS (Fig. 2C). Thus, unlike for OMKO1 and TIVP-H6, LPS-5 phage therapy did not show evidence for decreased LPS despite observed PsA resistance to LPS-5 (Table 2).

### Phage Therapy Does Not Change Bacterial Species Diversity

Although inhaled phage therapy reduced sputum bacterial load (Fig. 1A) and improved CF lung function (Fig. 1B), the possibility exists for administered phage(s) to alter non-target bacteria, including pathogens. To investigate this hypothesis, CF subject sputum bacteria was analyzed longitudinally. First, clinical laboratory sputum cultures showed no change in the number of CF pathogens post-phage therapy (Table 1). Next, we used longitudinal analyses of bacterial metagenomics from sputum samples obtained from subjects 2 through 9 over time, to gauge whether bacterial species differed relative to baseline (Fig. 4A). Data were examined using three separate species-richness metrics (i.e., Chao1, Shannon Evenness, and Simpson Index). Results showed no evidence that inhaled phage therapy led to changes in the presence of non-target bacterial species in the lung (Fig. 3A-D), and we concluded that the administration of phage(s) was neither beneficial nor costly to other detectable lung species.

**Figure 3.**
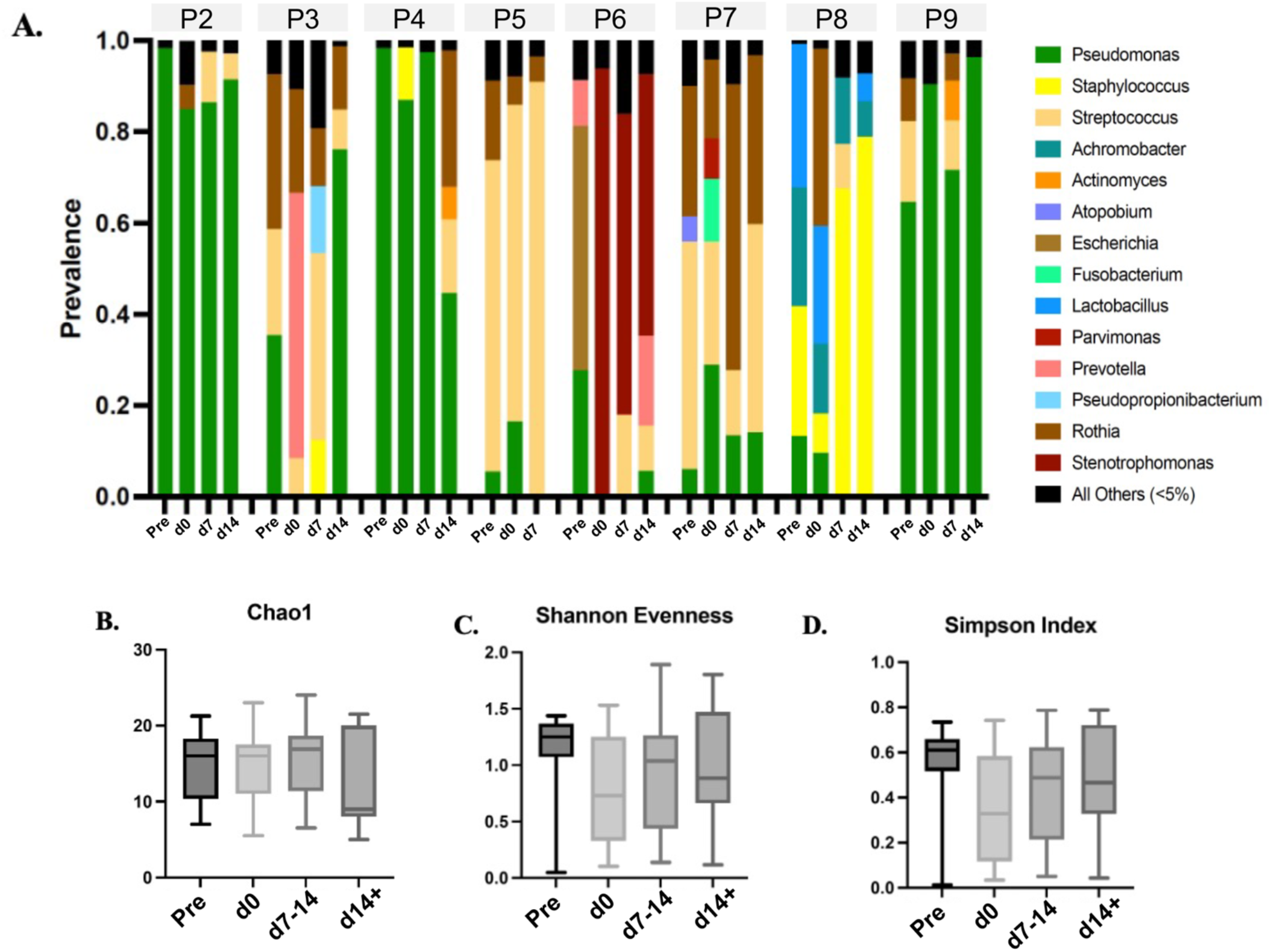
Effect of Phage Therapy on Sputum Microbiome. (**A**) Prevalence of bacteria genera (greater than 0.1%) in sputum samples for eight subjects as determined by sequence alignment. Comparison is made pre-phage (pre-) therapy, (tx), one week after phage therapy (1w), and post-phage (post-) therapy. Analysis includes average Chao-1 richness (**B**), average Shannon Evenness (**C**), and average Simpson Index (**D**) that shows no change from baseline for sputum from all subjects studied (Subjects #2-9).

## Discussion

Bacteriophages (phages) have been used clinically since shortly after their discovery in the early 20^th^ century. However, lack of standardization and published efficacy limited their use outside of eastern Europe until the emergence of antimicrobial resistance, which led to a resurgence of interest in phage therapy to treat MDR/PDR infections. Cystic fibrosis (CF) is a disease frequently complicated by recurrent, and increasingly MDR/PDR, pulmonary pathogens [12]. Thus, CF provides an opportunity to study the addition of phage therapy to standard care, and here we summarize our experience with adjunctive inhaled phage therapy to treat MDR/PDR PsA in nine CF adults that resulted in decreased sputum PsA and improved lung function, which may reflect the effects of phage-driven evolved trade-offs.

Phage therapy can be administered via different routes, which range from topical application to treat infected wounds, to intravenous (IV) delivery where phages are expected to reach the infected site/organ via the bloodstream. Inhaled phage therapy was first reported in 1959 [13], and, to date, we are only aware of two reports of inhaled phage therapy for PsA in two children with CF [14, 15]. For our consecutive cohort of 9 CF adults, nebulized phage therapy was chosen, in favor of IV delivery, to limit systemic phage exposure, with the added benefit that most individuals with CF have experience with inhaled medications. Nebulization should reduce gut microbiome exposure to therapeutic phages, minimize phage killing of commensal flora, and may decrease adaptive immune responses that target phages, which could limit future treatments with the same phages [16]. One limitation of this approach is that each case used an individual subject’s nebulizer, or one provided for them while inpatient. However, we confirmed that different non-mesh nebulizers had no impact on phage viability (data not shown), which suggests that nebulizer choice had no adverse effects on phage particles from two virus families (*Myoviridae* and *Podoviridae*). Importantly, sputum analysis showed that: 1) nebulized phage(s) decreased PsA CFU/ml (Fig. 1A), irrespective of nebulizer used, and 2) post-phage therapy sputum PsA showed evidence for bacterial resistance to treatment phages (Table 2), which suggests that inhaled phages affected their target MDR/PDR PsA identified in sputum. Therefore, this suggests that efficacy of inhaled phage therapy targeting PsA in CF adults may be generally robust, despite different nebulizer use.

The ability for inhaled phage therapy to decrease sputum PsA, especially for MDR/PDR bacteria, has the potential to translate into promising clinical applications, and our study outcome was similar in efficacy to inhaled or IV antibiotics targeting PsA in CF [17], although with a limited number of subjects. Thus, our approach robustly reduced CFU despite each subject in the cohort being infected with a different MDR/PDR PsA strain, and all having recently completed antibiotics, or concurrently receiving antibiotics (Table 1). Nevertheless, decreased CFU does not always correlate with clinical improvement [18]. It was therefore notable that all nine subjects in this cohort had improved lung function (Fig. 1B), albeit to different degrees, which suggests the potential for generalized clinical relevance of inhaled phage therapy in CF.

A longstanding concern surrounding phage therapy is that target bacteria will (perhaps inevitably) evolve resistance to administered phages, which is an inherent weakness of phage therapy [4]. Therefore, beyond demonstrating decreased CFU and pulmonary improvement due to inhaled phage therapy, our study sought to test the hypothesis that evolved phage resistance could be leveraged as a benefit, rather than constituting a weakness. We predicted that specific phages should select for favorable reductions (trade-offs) in PsA virulence if the target bacteria evolved phage resistance. We previously used this strategy to treat a MDR PsA aortic-graft infection with phage OMKO1 where susceptible PsA evolved resistance to this phage along with re-sensitivity to effluxed antibiotics *in vitro*, which was presumably caused by PsA evolving phage resistance by reducing (or eliminating) efflux-pumps [8]. Similarly, in this CF cohort, post-phage therapy PsA isolates were resistant to phages (Table 2), which resulted in increased antibiotic sensitivity (targeting efflux pumps with phage OMKO1; Fig. 2A). Because OMKO1 increased antibiotic sensitivity, clinicians were able to use antibiotics that were previously unavailable because of MDR or PDR PsA resistance. There was similar evidence for decreased virulence in post-phage therapy PsA isolates targeting TIVP with phage TIVP-H6 (Fig. 2B). However, despite evidence that PsA sputum isolates treated with phage LPS-5 became resistant to this phage (Table 2), there was no measurable difference in LPS in post-phage therapy PsA supernatants (Fig. 2C). Potential explanations for this observation include: 1) PsA evolved resistance to LPS-5 in receptor-independent mechanism(s), 2) direct measurement of LPS does not reflect a potential LPS-5 phage driven trade-off (e.g., biofilm, cell permeability or viability changes), which is an area of ongoing investigation, and/or 3) phage cocktail approach was sufficient to select for decreased PsA virulence in most (seven of nine) subjects (Figs. 2A & 2B), relative to effects of phage LPS-5 selection. The latter explanation is consistent with observed resistance of *Escherichia coli* to a specific phage via efflux pump mutations, which are less costly for bacterial growth than changes in LPS [19]. An important limitation is that, while a small number of PsA isolates chosen at random from subject sputum was sufficient to make phage therapy decisions, we cannot definitively confirm whether post-phage therapy isolates were truly direct descendants of the original pre-phage therapy PsA. The observed resistance of post-phage therapy PsA isolates to specific phages administered was consistent with our proposed trade off mechanism. However, it is plausible that PsA resistance driven by one phage was sufficient to achieve reduced virulence when patients received cocktails. Overall, our data indicated that irrespective of PsA lineage, phage therapy predictably affected CF PsA sputum communities, and most patients yielded PsA isolates with virulence trade-offs, which may have contributed to improved lung function.

While PsA is currently the most prevalent pathogen observed in CF adult sputum, polymicrobial infections (e.g., *Staphylococcus aureus*) are common, and evident by studies investigating the CF lung microbiome utilizing culture-independent techniques that detect multiple pathogens [20, 21]. Given the potential for phage therapy to decrease PsA while increasing the opportunity for competitively inferior pathogens to thrive, a crucial aspect of our study was to investigate how phage therapy might alter the CF lung microbiome, which was examined using longitudinal analysis of metagenomics data for each subject. We found no evidence that phage therapy altered species compositions in the CF lung microbiome (Fig. 3). These observations were consistent with clinical laboratory analyses of post-phage sputum samples, which also showed no changes in the number of pathogens (Table 1).

Many aspects of our study addressed possible safety concerns for phage therapy, which does not have U.S. FDA approval for general use. This study used environmentally sourced phages that were unaltered by genetic manipulation, which is analogous to approved applications generally recognized as safe (GRAS) by the FDA. Safety was addressed by restricting administered phages to those with strictly lytic replication cycles, thus avoiding temperate phages capable of host lysogeny (stable integration into the host bacterial chromosome, allowing phage inheritance by daughter cells). Each phage was genome sequenced and screened for presence of lysogeny genes (data not shown) to further ensure phages were strictly lytic. In addition, purification techniques were employed to minimize endotoxin levels in phage doses to ensure these were below FDA requirements [< 5 endotoxin units (EU)/kg/h]. Because pre-phage CF sputum PsA CFU was ∼3 × 10^8^ CFU/ml, a phage concentration was chosen to exceed bacterial density by at least an order of magnitude [i.e., multiplicity of infection (MOI) ≈ 10]. Although this method was successful in the current study, a unique benefit of lytic phage therapy is viral self-amplification within their host bacteria, suggesting that relatively lower concentrations, perhaps MOI < 1, could be considered with future inhaled phage therapies. However, we note that the current high MOI approach may leverage a benefit that is analogous to ‘migration pressure’ in a biological system, whereby the persistent influx of a single immigrant genotype (here, the wildtype phage) can numerically overwhelm the adaptive potential of a resident population (the phages inhaled on prior days that could become locally established) [22, 23]. Therefore, multiple daily doses containing large numbers of wildtype phage may cause phage evolution within the CF lung to ‘stall out’, because the wildtype perpetually dominates the population during treatment, minimizing the adaptive potential of new genetic variants, including ability to coevolve with resident PsA bacteria. This migration pressure and reduced potency for coevolution should foster greater accuracy when predicting evolved trade-offs in target bacteria and is consistent with intentions to minimize duration of phage therapy, reduce systemic effects, and limit human adaptive immune responses.

Phage cocktails are often assumed, but not confirmed, as the superior approach to phage therapy. Although phage cocktails can combine viruses to usefully kill a greater diversity (broader range) of host strains, relative to each phage alone, this popular approach has certain limitations: 1) different viruses co-infecting the same bacterial cell can suffer reduced reproductive fitness due to phage competition for limited intracellular resources [24, 25]; 2) cocktails can select for unanticipated phage-resistance mutations compared to single phages [26, 27], creating concerns for unexpected evolved changes in bacterial virulence and/or pathogenicity; 3) cocktails may stimulate greater human adaptive immune responses compared with single phages [28]; and 4) cocktails have the potential to select for pan-phage-resistance in bacteria, which more quickly depletes libraries of potentially useful phages [4]. We observed clinical efficacy with individual phages TIVP-H6 and LPS-5, although only TIVP-H6 showed evidence for the predicted virulence trade-off (Table 2; Fig 2B). This suggests that single and more likely, single-sequential phage therapy may be effective to address CF pulmonary infections. This approach is attractive owing to the need to treat PsA, or other bacteria, for prolonged durations because of the inability to fully eradicate bacteria within CF bronchiectatic lungs. In addition, a single-sequential approach could be deployed such that alternating phages would target different bacterial surface receptors to drive specific changes in PsA virulence post-phage therapy. This serial approach would leverage the same phage discovery platform that we used to discover OMKO1, LPS-5, and TIVP-H6, to search for PsA phages that target additional virulence factors (e.g., secretion systems, flagella, and siderophores). Single-sequential nebulized phage therapy may also limit adaptive immune responses, although this requires more rigorous study. This approach would also allow for the potential use of higher phage concentrations because a fixed limitation in phage therapy is endotoxin concentration per each phage preparation. Some naturally occurring phages are capable of broadly killing genotypes of host bacteria, including PsA [29], and a general goal of phage biotechnology is often to identify or engineer viruses with broad host range [30]. Vital data that relate to the above limitations are being studied in an investigator-initiated clinical trial at Yale (NCT04684641) and Armata Pharmaceutical (NCT04596319) and BiomX Inc (NCT05010577) trials, which examine the safety and potential efficacy of inhaled phage therapy using single versus cocktail based approaches in CF, respectively.

In summary, this study represents the largest cohort of compassionate cases of adjunctive nebulized phage therapy for MDR/PDR PsA in PwCF. While the number of subjects is limited, nebulization was well-tolerated, and results showed that a personalized phage therapy approach selected phages that effectively target and kill sputum MDR/PDR PsA. In addition to this bacterial killing, this phage therapy approach confirmed that using specific phages that drive trade-offs in PsA should result in decreased PsA antibiotic resistance and virulence, because certain phages exploit host receptors that exert selection pressure for target PsA to evolve phage resistance that coincides with clinically useful reductions in virulence. Thus, in a chronic lung disease such as CF, inhaled phage therapy may offer a novel therapeutic option, which is already being studied in ongoing clinical trials. In addition, this approach may be relevant to treat additional MDR/PDR ESKAPE pathogens as the complexity, and mortality, from AMR infections continues to increase [1].

## Online Methods

### Identification of patients suitable for phage therapy

Between 12 December 2017 and 26 May 2019, we received unsolicited requests from physicians and/or their CF patients for assistance to treat multi-drug resistant (MDR) or pan-drug resistant (PDR) sputum pathogens. Most samples were for *P. aeruginosa* (PsA) infections, but these requests also included a variety of other pathogens. For each case, our program (phage biologists, CF, and Infectious Disease physicians) reviewed the clinical history, antibiotic treatment regimens, and immunologic status of each patient. Spontaneously expectorated sputum was sent to what is now Yale’s Center for Phage Biology & Therapy laboratory for microbiological characterization. Here we describe our experience with the first nine CF patients treated with nebulized phage therapy for PsA (Table 1). PsA was isolated from sputum (see below) and susceptibility to three phages [OMKO1 (targeting Mex efflux pumps), LPS-5 (targeting LPS), TIVP-H6 (targeting type IV pilus)] was determined (Table 2). Each phage therapy protocol was submitted as an individual FDA emergency investigational drug (eIND) application, each subject provided consent, and each eIND protocol was IRB reviewed at the institution where patients received phage therapy. Subjects were treated at their CF program (n = 4) or traveled to Yale (n = 5). The Yale Center for Phage Biology & Therapy prepared and provided phages for all subjects. Phage therapy was delivered by nebulization. This approach was chosen because inhalation is generally well-tolerated in CF, as many therapies (e.g., mucolytics and antibiotics) are nebulized, and nebulization has less potential to result in systemic toxicity compared to other routes of delivery, although this has not been rigorously investigated to date. For each patient, phage therapy occurred during, or immediately after, antibiotic treatment for MDR/PDR PsA. All patients received an initial phage therapy dose (“test dose”) in either an inpatient or outpatient clinical setting where they were monitored after nebulization, using a non-mesh jet nebulizer (inpatient: provided by the hospital; outpatient: patient’s own nebulizer), to ensure that inhalation was well tolerated. All patients tolerated this “test dose” without any evidence for respiratory distress or increased cough. For inpatients (n = 3), phage therapy was continued twice daily, while outpatients (n = 6) received phage therapy once daily. Three subjects transitioned from inpatient to outpatient. Total duration of phage therapy was for 7 to 10 days. For outpatients with initial precent predicted forced expiratory volume in one second (ppFEV_1_) <30%, clinical follow-up occurred within 72 hours of treatment. After phage therapy, sputum samples were collected and processed (see below), and spirometry was performed at approximately 21 to 28 days after phage therapy.

### Sputum processing

All sputum samples were spontaneously expectorated. Parallel sputum samples were sent to clinical laboratories for standard microbiology analysis. In Yale’s Center for Phage Biology & Therapy laboratory sputum collected for research was weighed and combined with 1 ml PBS [with 10 mM magnesium sulfate (MgSO_4_)] per gram and manually homogenized with a syringe and 16-gauge needle [31]. Samples were then immediately stored in DNA/RNA Shield (Zymo Research) per manufacturer protocol prior to metagenomic sequencing (see below).

### Bacterial isolation, culture, and quantification

Bacteria were isolated by direct plating 10 μl of expectorated sputum homogenate on PsA isolation agar. This sample was used to measure PsA colony-forming units (CFU) and to isolate phenotypically distinct strains for further characterization (n = 1-5 per sputum sample). As previously described [8, 9], CFU were estimated for a bacterial strain *via* dilution series of stationary-phase cultures, followed by plating on Tryptic Soy Broth (TSB) agar, and countable CFU were used to calculate cell densities (CFU/ml of expectorated sputum). The Wilcoxon signed rank t test for paired samples was used for statistical analysis. For subsequent experiments, all bacteria were grown from frozen stock on 1.5% agar made from TSB, by streaking for single colonies that were grown for 24 hours at 37° C. Individual colonies were used to initiate cultures grown overnight in TSB with shaking (150 rpm) incubation at 37° C. Metagenomic sequencing was conducted using deep sequencing data obtained on the Illumina HiSeq4000 platform to analyze genomes extracted from uncultivated bacterial species present in communities within patient sputum samples. Metagenomic sequencing results were analyzed to examine longitudinal changes in alpha-diversity (species richness) indices across samples for each patient using three different metrics: 1) Chao1 diversity index is a richness metric that especially considered potentially missing (‘rare’) species, 2) Simpson diversity index was used to account for presence of specific dominant species, aside from overall abundance, and 3) Shannon diversity index was used to account for abundance and evenness of species across time.

### Determination of phage(s) used in treatment

Candidate phages were tested on patient sputum PsA. Dilutions of a phage lysate were tested for ability to form plaques on the test strain, relative to plaquing ability on a permissive PsA laboratory strain, either PAO1 or PA14 [32]. Candidate phages for therapy were subjected to whole-genome sequencing; DNA was extracted using the Norgen Biotek phage DNA isolation kit, and sequencing library was prepared using the Nextera DNA Flex library preparation kit and sequenced on an Illumina NextSeq system, yielding 150-bp paired-end reads. Results were analyzed using BLASTn [33], to avoid temperate, lysogenic phages (i.e., those able to lysogenize their bacterial host) and to ensure that strictly, lytic phages did not contain potentially harmful genes, such as toxins.

### Phage preparation

Phage lysates were prepared by growing laboratory PsA PAO1 or PA14 to exponential phase in TSB [30g of TSB powder (non-animal origin)] in 1 L diH20 (per manufacturer protocol, Sigma Aldrich). Phage(s) were then mixed with the bacteria at multiplicity of infection (MOI; ratio of phage particles to bacterial cells) of ∼0.01 and incubated at 37° C with shaking (100 rpm). After 6 hours, cultures were centrifuged and filtered (pore size: 0.22μm) to obtain a cell-free phage lysate. Lysate was then concentrated with Centricon small pore concentrators (100 kDa MWCO) and dialyzed in 1000x volume PBS with MgSO_4_. Endotoxin concentration was then determined with Hyglos EndoNext kits (bioMerieux Inc., Durham, NC). Per FDA recommendations, USP 71 testing was completed by a third-party laboratory (Accugen Laboratories) on all phage preparations that were used for phage therapy. The final preparation was then diluted in PBS with MgSO_4_ to a concentration of 1.0 × 10^10^ PFU/ml for individual phages. If phages were used in a cocktail (2 or more phages), total phage concentration remained ≤1.0 × 10^10^ PFU (e.g., 2 phages ∼5.0 × 10^9^; 3 phages ∼3.33 × 10^9^). Final phage concentration was diluted into 3 ml PBS for nebulization. If shipping was required, phages were shipped on ice and kept refrigerated prior to use.

### Bacterial supernatants

Pre- and post-phage therapy individual colony bacteria cultures from sputum were grown from frozen stock for 12 to 48 hours in lysogeny broth (LB; 10 g tryptone, 5 g yeast extract, and 10 g NaCl per L of distilled water) or TSB with shaking (150 rpm) incubation at 37° C and optical densities at wavelength λ = 600 nm (OD_600_) were estimated *via* spectrophotometry. Each culture was diluted in LB or TSB to OD_600_ = 0.4 and then centrifuged for 15 minutes at 5000 xg and supernatants passed through a 0.2 μm filter to remove residual bacterial cells.

### Lipopolysaccharide (LPS) measurements

Endotoxin/LPS was quantified in bacterial culture cell-free supernatants *via* ENDONEXT kits (bioMerieux Inc., Durham, NC) *per* manufacturer protocol. LPS EU/ml was measured (corrected for CFU/ml), in pre- and post-LPS phage cell-free PsA supernatants from Subjects 4 and 5, individuals who received LPS-5 single phage therapy. Comparison was made to *Pseudomonas* laboratory strain PA14, which is used to amplify phage LPS-5. Statistical analysis used Welch’s t test.

### Bacteria attachment assay

PsA cultures were grown 12-24 hours in TSB (15 g of BD™ Bacto™ Tryptic Soy Broth, BD™ Soybean-Casein Digest Medium, in 500 ml of purified water) then washed with MEM (Gibco) and diluted to OD_600_ of 0.1. Normalized bacteria were then added to an immortalized cystic fibrosis ΔF508/ΔF508 bronchial epithelial (CFBE41o-) cell line [generously provided by Dr. J. Bomberger (University of Pittsburgh)] [34]. CFBE41o-cells were cultured at 37° C and 5% CO_2_ in minimal essential medium (MEM) (Gibco) with 10% fetal bovine serum (FBS; Gemini Bio-Products), supplemented with 2 mM l-glutamine, 5 U/ml penicillin, and 5 μg/ml streptomycin (Sigma); and 0.5 μg/ml plasmocin prophylactic (InvivoGen). Cells were seeded at confluence on transwell Falcon^®^ Permeable Support and differentiated at air-liquid interface for 7 days prior to use. Normalized bacteria and CFBE41o-were co-cultured at 37° C for 1 hour, as previously described [35]. The apical medium was removed, and bronchial cells washed thoroughly with MEM. Attached bacteria were dispersed with MEM containing 0.1% Triton x-100 solution, serially diluted, and streaked on LB agar plates [12.5 grams of LB (Miller’s LB Broth Base™) and 15 g agar in 500 ml of purified water] to determine CFU counts. Bacteria from different *Pseudomonas* laboratory strains were studied for comparison: PA103 wildtype and PA103 TIVP mutants (PA103ΔpilA and PA103ΔpilQ). PsA isolates pre- and post-TIVP-H6 phage from Subject 2, the only individual who received TIVP-H6 single phage therapy, were studied in this attachment assay. Results are expressed as CFU percentage of the original inoculum. Statistical analysis used Welch’s t test.

### Spirometry

Forced expiratory volume in one second (FEV_1_) was obtained from each patient by spirometry per each CF clinic’s protocol. All clinics used American Thoracic Society standards of acceptability and repeatability. The Wilcoxon signed rank t test for paired samples was used for statistical analysis.

### Statistics

List things done in Prism, List things done in R. What was done for each methods. All statistical analysis was reviewed by Dr. Shabanova. Need software. Need references for analysis methods of sputum. (Doug Conrad metagenomics and sub-analysis.

## Data Availability

All data produced in the present study are available upon reasonable request to the authors

## Acknowledgements

We thank Dr. J. Bomberger (University of Pittsburgh) for assistance with attachment assay; Dr. P. Caudill (Yale University) for kindly ensuring phage manufacturing compliance; A. Hummel (Yale University) and the Yale Center for Clinical Investigation for assistance with the eIND process; M. Baranoski at the Yale University Institutional Review Board; C. Fiore (FDA) for invaluable advice and assistance in these cases.

